# Pediatric ARDS phenotypes in critical COVID-19: implications for therapies and outcomes

**DOI:** 10.1101/2022.06.08.22276125

**Authors:** Jesús Domínguez-Rojas, Yesica Luna-Delgado, Patrick Caqui-Vilca, Carlos Martel-Ramírez, Miguel Quispe-Chipana, Mario Cruz-Arpi, Noé Atamari-Anahui, Cleotilde Mireya Muñoz Ramírez, Gaudi Quispe Flores, Mariela Tello-Pezo, Pablo Cruces, Pablo Vásquez-Hoyos, Franco Díaz

**Affiliations:** Hospital Nacional Edgardo Rebagliati Martins, Departamento de Pediatría, Lima, Perú; Hospital Nacional de Emergencias de Villa El Salvador. División de Cuidados Críticos Pediátricos, Lima, Perú; LARed Network, Red Colaborativa Pediátrica de Latinoamérica; Hospital Nacional Regional de Cusco, División de Cuidados Críticos Pediátricos, Perú; Hospital Nacional Hipólito Unanue, Lima, División de Cuidados Críticos Pediátricos, Lima, Perú; Universidad San Ignacio de Loyola, Vicerrectorado de Investigación, Unidad de Investigación para la Generación y Síntesis de Evidencias en Salud, Lima, Perú; Instituto Nacional de Pediatría, División de Cuidados Críticos Pediátricos, Ciudad de México; Instituto Nacional de Salud del Niño San Borja. Departamento de Pediatría, Lima, Perú; Hospital El Carmen de Maipú, Unidad de Cuidados Intensivos Pediátricos, Maipú, Chile; Facultad de Ciencias de la Vida, Universidad Andres Bello, Santiago Chile; Departamento de Pediatría, Fundación Universitaria de Ciencias de la Salud, Bogotá, Colombia; Departamento de Pediatría, Universidad Nacional de Colombia, Bogotá, Colombia; Escuela de Medicina, Universidad Finis Terrae, Santiago, Chile

**Keywords:** PARDS, COVID-19, MIS-C, Pulmonary mechanics, Driving pressure

## Abstract

**Purpose:** to describe lung mechanics in Pediatric Acute Respiratory Disease Syndrome (PARDS) associated with COVID-19. We hypothesize two phenotypes according to respiratory system mechanics and clinical diagnosis.

**Methods:** a concurrent multicenter observational study was performed, analyzing clinical variables and pulmonary mechanics of PARDS associated with COVID-19 in 4 Pediatric intensive care units (PICUs) of Perú. Subgroup analysis included PARDS associated with multisystem inflammatory syndrome in children (MIS-C), MIS-PARDS, and PARDS with COVID-19 primary respiratory infection, C-PARDS. In addition, receiver operator curve analysis (ROC) for mortality was performed.

**Results:** 30 patients were included. Age was 7.5(4-11) years, 60% male, and mortality 23%. 47% corresponded to MIS-PARDS and 53% to C-PARDS phenotypes. C-PARDS had positive RT-PCR in 67% and MIS-PARDS none (p<0.001). C-PARDS group had more profound hypoxemia (P/Fratio<100, 86%vs38%,p<0.01) and higher driving-pressure (DP) [14(10-22)vs10(10-12)cmH_2_O], and lower compliance of the respiratory system (C_RS_)[0.5(0.3-0.6)vs 0.7(0.6-0.8)ml/kg/cmH_2_O] compared to MIS-PARDS (all p<0.05). ROC-analysis for mortality showed that DP had the best performance [AUC 0.91(95%CI0.81-1.00), with the best cut-point of 15 cmH_2_O (100% sensitivity and 87% of specificity). Mortality in C-PARDS was 38% and 7% in MIS-PARDS(p=0.09). MV free-days were 12(0-23) in C-PARDS and 23(21-25) in MIS-PARDS(p=0.02)

**Conclusion:** critical pediatric COVID-19 is heterogeneous in children. COVID-19 PARDS had two phenotypes with distinctive pulmonary mechanics features. Characteristics of C-PARDS are like a classic primary PARDS, while a decoupling between compliance and hypoxemia was more frequent in MIS-PARDS. In addition, C-PARDS had fewer MV free-days. DP ≥ 15 cmH2O had the best performance of the quasi-static calculations to discriminate for mortality. Standardized pulmonary mechanics measurements in PARDS might reveal essential information to tailor the ventilatory strategy in pediatric critical COVID-19.

**‘Take-home message’:**

- PARDS associated with COVID-19 have two different phenotypes based on clinical diagnosis and pulmonary mechanics.
- C-PARDS group was characterized as a classic moderate to severe primary ARDS. A decoupling between compliance and hypoxemia was more frequent in MIS-PARDS. Regarding outcomes, C-PARDS had less VFD and a trend toward higher mortality.
- Data from the quasi-static calculations were associated with mortality; DP≥ 15 cmH2O was the best discriminator.
- Standardized pulmonary mechanics measurements in PARDS might reveal essential information to tailor the ventilatory strategy, characterizing different phenotypes and parameters associated with outcomes.

**Tweet:** Lung mechanics help to differentiate two different phenotypes in PARDS associated with COVID-19. C-PARDS associated with respiratory infection, and MIS-PARDS, associated with MIS-C. Also, lung mechanics variables were associated with mortality, being DP ≥ 15 cmH_2_O the best discriminator.

## BACKGROUND

Respiratory failure has been the leading cause of hospital admission and death during the COVID-19 pandemic. Severe hypoxemia due to pneumonia and acute respiratory distress syndrome (ARDS) is a frequent cause of admission to intensive care for advanced respiratory support^1^. Since the first cohorts’ description in China and Europe, many authors reported discrepancies between the severity of the oxygenation and the relatively spared pulmonary mechanics in a subgroup of patients^2,3^

In addition, atypical lung imaging in chest computed tomography and histopathology with lung microvascular involvement raises questions about whether the underlying pathophysiology in COVID-19 is like that of ARDS in other etiologies^2,4,5,6^. Thus, a new entity called C-ARDS (COVID-19 associated ARDS) is proposed by some researchers^7-9^.

Pediatric COVID-19 critical illness is heterogeneous and infrequent^10-12^. The most common causes of admission to the pediatric intensive care unit (PICU) are respiratory failure and multisystem inflammatory syndrome in children (MIS-C),^10-15^. Nonetheless, cases of non-cardiopulmonary involvement are frequent^14-16^. While the low morbidity and mortality in the general pediatric population is reassuring, there is a significant gap in knowledge in identifying high-risk subgroups, such as those who develop pediatric ARDS (PARDS) or multiorgan failure. ^11,12,16^

Invasive mechanical ventilation (MV) for pediatric COVID-19 at PICU has been reported between 30 to 70% in different cohorts^11,13,15,17-19^. Surprisingly, specific information on PARDS related to COVID-19 is scarce. The heterogeneous nature of lung involvement was initially described as distinctive phenotypes in adult C-ARDS based on pulmonary mechanics, type L and type H^2^. Although still controversial, different phenotypes might have implications for therapy and outcomes^20,21^. This principle also applies to respiratory failure and PARDS in the general PICU population. A better description of PARDS characteristics is urgently needed to improve guidelines and recommendations and ultimately improve outcomes for critically ill children.

We hypothesize that two different phenotypes can be observed in critically ill children with PARDS associated with COVID-19: 1) PARDS in patients diagnosed with MIS-C (MIS-PARDS), with almost normal compliance, low driving pressure, and extrapulmonary organ failure; 2) PARDS with severe COVID-19 pneumonia (C-PARDS), with low compliance, profound hypoxemia, and less frequent multiorgan failure. This study aimed to describe lung mechanics in critically ill children with PARDS due to COVID-19, analyzing sub-groups of MIS-C and C-PARDS and outcomes.

## PATIENTS AND METHODS

A concurrent observational study was conducted in four PICU’s of pediatric referral hospitals in Perú: Hospital Nacional Hipolito Unanue, Hospital de Emergencias de Villa El Salvador, Hospital Regional del Cusco, Hospital Edgardo Rebagliati Martins. Institutional review boards’ approval for data collection was obtained at each site, waiving informed consent

The study included patients between 1 month and 17 years of age from the PICU Registry cohort of COVID-19 admitted between April 1 and August 31, 2021. Briefly, this observational registry records the treatment and the management of critical COVID-19 patients admitted to participating centers. Deidentified data were gathered from administrative and clinical databases for benchmarking, including demographics, clinical and physiological parameters, therapeutic interventions, and outcomes. Critical COVID-19 definition included patients with SARS-CoV-2 rt-PCR in respiratory airways or antibody profile compatible with acute COVID-19 infection (IgG (+)) and CT scan with characteristics of acute COVID-19 infection. In addition, patients who met the case definition for MIS-C were also included, even lacking a confirmed microbiological diagnosis of SARS-CoV-2 infection, to include all the causes of pediatric critical COVID-19, requiring PICU admission, as previously defined. ^11,12,22,23^

Patients with a clinical and microbiological profile of SARS-CoV-2 infection at PICU admission, receiving invasive MV, and meeting PALICC criteria for PARDS^24^, were included in the analysis. Pneumonia severity was based on PAHO definitions^25^. Patients were excluded if they had uncorrected congenital heart, pre-existing lung or airway disease, chronic respiratory failure requiring long–term MV and tracheostomy, severe left ventricular dysfunction failure with ejection fraction less than 30%^26^ and COVID-19 with primary neurological involvement. In addition, patients with spontaneous breathing effort and endotracheal tube air leak were excluded due to possible interference with data acquisition and unreliable quasi-static lung mechanics measurements and calculations.

During the first 72 hrs after initiation of MV, all patients were screened at 8 AM and 8 PM. PARDS severity was determined following PALICC criteria, according to the oxygenation index, classified as mild (4 to <8), moderate (8 to <16), and severe (≥ 16).^24^ When hypoxemia was the lowest, all the parameters were recorded. Organ dysfunction was assessed by the treating physician based on definitions by the international pediatric sepsis consensus conference^27^, and multiorgan dysfunction was defined as ≥ 3 organ dysfunctions. Vasoactive support was quantified through Vasoactive-Inotropic Score (VIS)^28^. Cardiac function was evaluated in all patients with transthoracic echocardiography performed by an experienced clinician, defining cardiac dysfunction as any alteration in systolic and diastolic function, and severe dysfunction was defined as ejection fraction less than 40%^26, 29^. In assessing organ dysfunction, we consider the worst value in the first 72 h after admission. Analyzed outcomes were duration of MV, Ventilator-free days at day 30 (VFD), Length of PICU stay, length of hospital stay, and PICU mortality.

Patients were classified into the MIS-C (MIS-PARDS) and COVID-19 pneumonia (C-PARDS) phenotypes. All patients fulfilling MIS-C diagnostic criteria (World Health Organization (WHO), Royal College of Paediatrics and Child Health, or Centers for Disease Control and Prevention) were classified as MIS-PARDS^30-32^. C-PARDS group consisted of PARDS patients fulfilling COVID-19 pneumonia according to the modified case definitions of the World Health Organization (WHO).^33^ Supplementary table 1 shows detailed patient classifications according to diagnostic tests.

Lung mechanics were measured in a volume control mode as previously described.^34,35^ Ventilator parameters [peak inspiratory pressure (PIP), plateau pressure (Pplat), PEEP, expiratory VT (VTE), and inspiratory time (IT), and arterial blood gases were registered, and PaO_2_/FiO_2_ ratio and Oxygenation Index (OI) were calculated. The components of working pressure, resistive (PIP – Pplat) and elastic (driving pressure, ΔP = Pplat–PEEP), were calculated for each subject. Respiratory system compliance (C_RS_, mL·cmH2O^-1^·kg^-1^) was calculated according to the standard equation.

### Statistical analysis

The data obtained were entered in a database on Microsoft® Excel (version for Windows 2016), reviewed, cleaned, and analyzed in STATA v.16 (StataCorp LP, Texas, USA). Frequencies and percentages were used to describe the categorical variables, while median and interquartile range (IQR) were used for quantitative data since the assumption of normality was not met. In addition, a bivariate analysis was performed between MIS-PARDS and C-PARDS and for survivors and non-survivors, using Fisher’s exact test for categorical variables and the Mann-Whitney U test for quantitative variables. A p-value less than 0.05 was considered statistically significant. Finally, receiver operator characteristic (ROC) curves were built for DP, C_RS,_ and Pplat, to evaluate their accuracy as discriminators for mortality as an outcome.

## RESULTS

During the study period, 123 critically ill COVID-19 children were admitted to the participating units, 30 with PARDS diagnosis, and all of them met the selection criteria for the analysis (Figure 1). The median age was 7.5 years (4-11), 63% were intubated before PICU admission, 50% of the patients suffered moderate PARDS, 30% severe, and 30% developed multiorgan failure. PICU median length of stay was seven days (5-9), and mortality was 23% (Table 1). The primary cause of death was refractory shock and multiorgan failure.

**Figure. 1.**
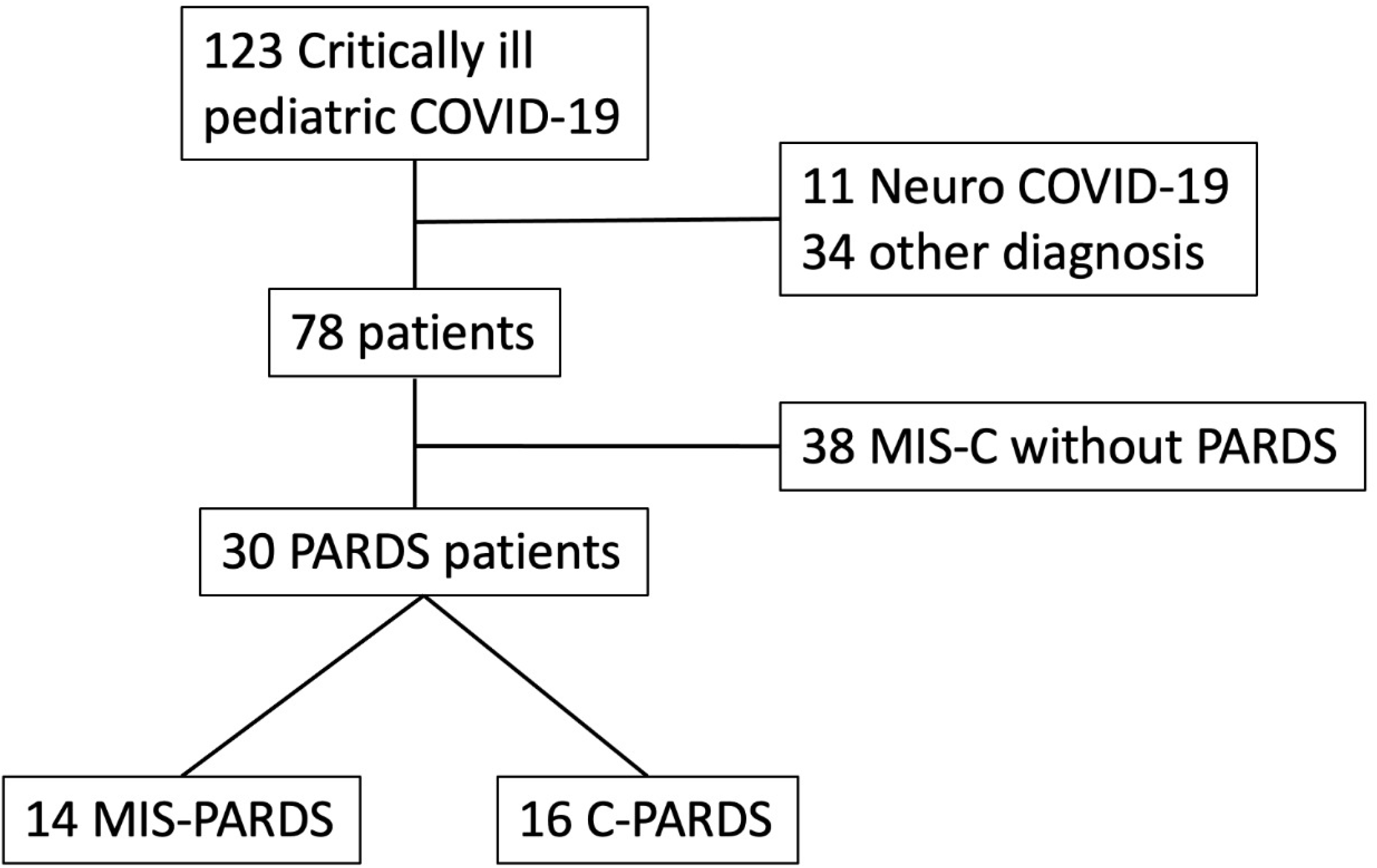
Study patient flow diagram. (PARDS: Pediatric Acute Respiratory Distress Syndrome; C-PARDS, PARDS associated with COVID-19 pneumonia; MIS-PARDS, PARDS associated with multisystem inflammatory syndrome in children)

**Table 1.**
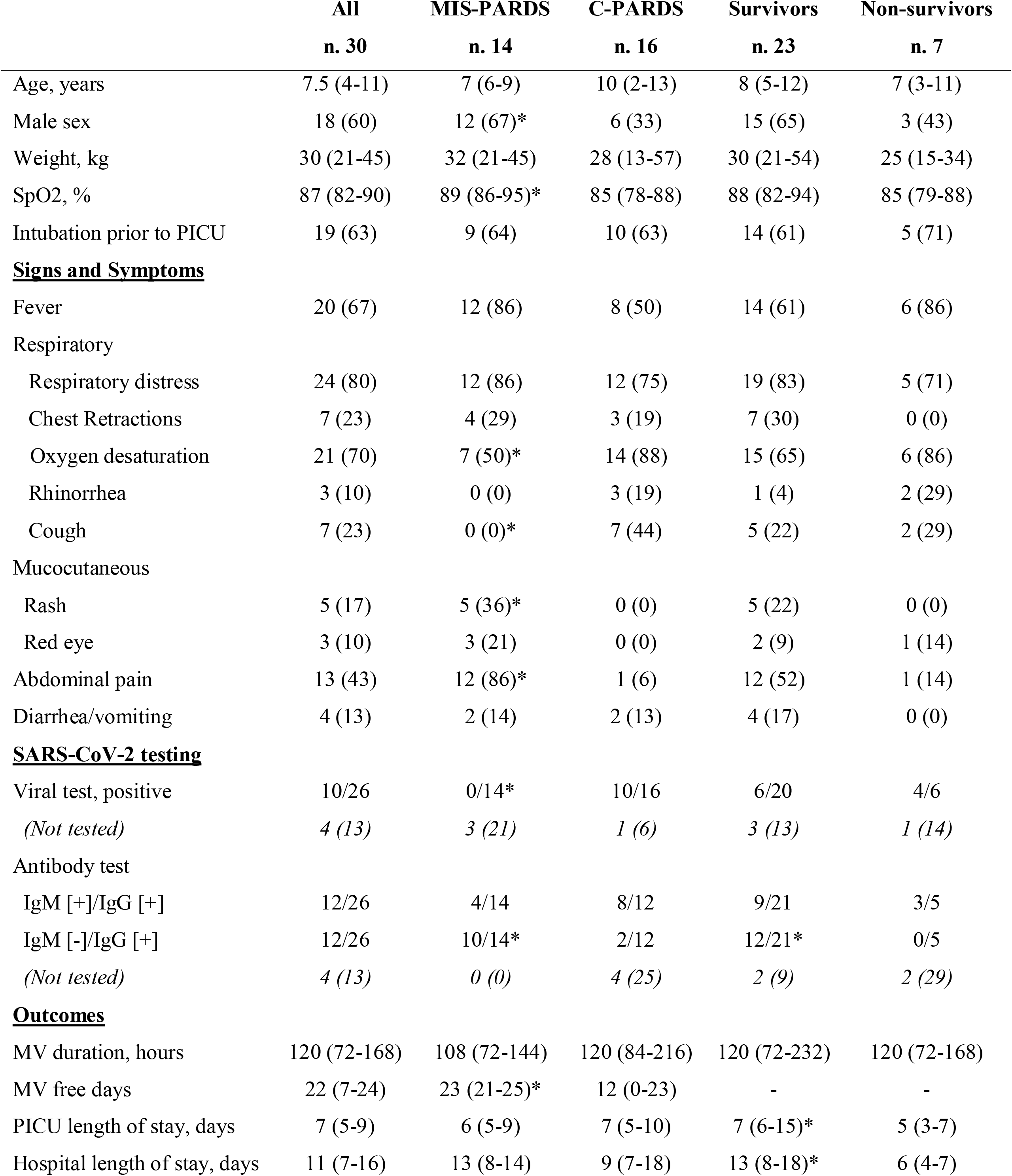
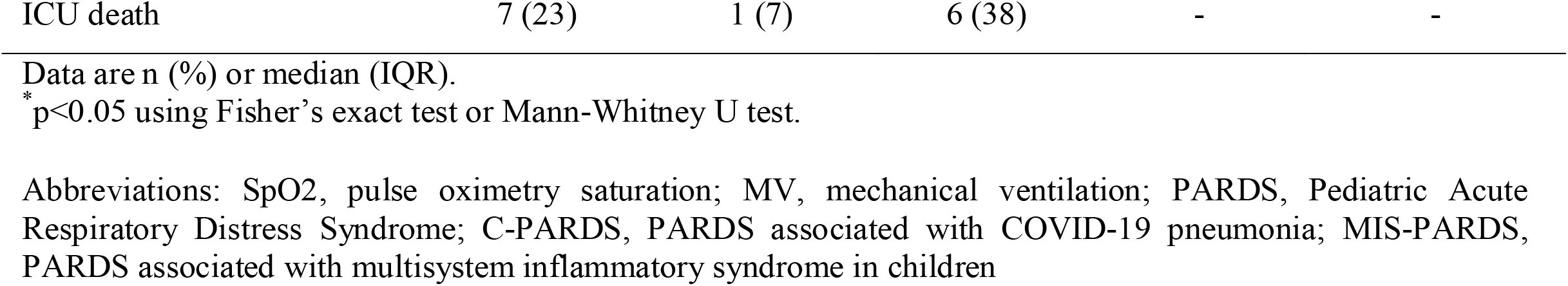
Demographic and clinical characteristics of pediatric respiratory distress syndrome associated with COVID-19 according to the clinical phenotypes and outcome.

The most frequent clinical signs and symptoms were dyspnea (77%), fever (67%), and abdominal pain (43%). Regarding the COVID-19 diagnosis, RT-PCR was positive in 38% of PARDS patients, and serology tests showed that 12/26 patients were IgM/IgG positive and 12/26 were IgM negative/IgG positive. Only one case was positive for IgG and RT-PCR. Viral co-infection and bacterial superinfection were ruled out with PCR for viral respiratory pathogens and blood and tracheal secretion cultures.

In 5 cases, left ventricular dysfunction was observed in echocardiograms, but none were severe (Supplementary Table 2). Coronary aneurysms were reported in two patients. The most frequent vasopressors were epinephrine (67%) and norepinephrine (57%), and VIS was 22 (12 to 50) (Table 2).

**Table 2.**
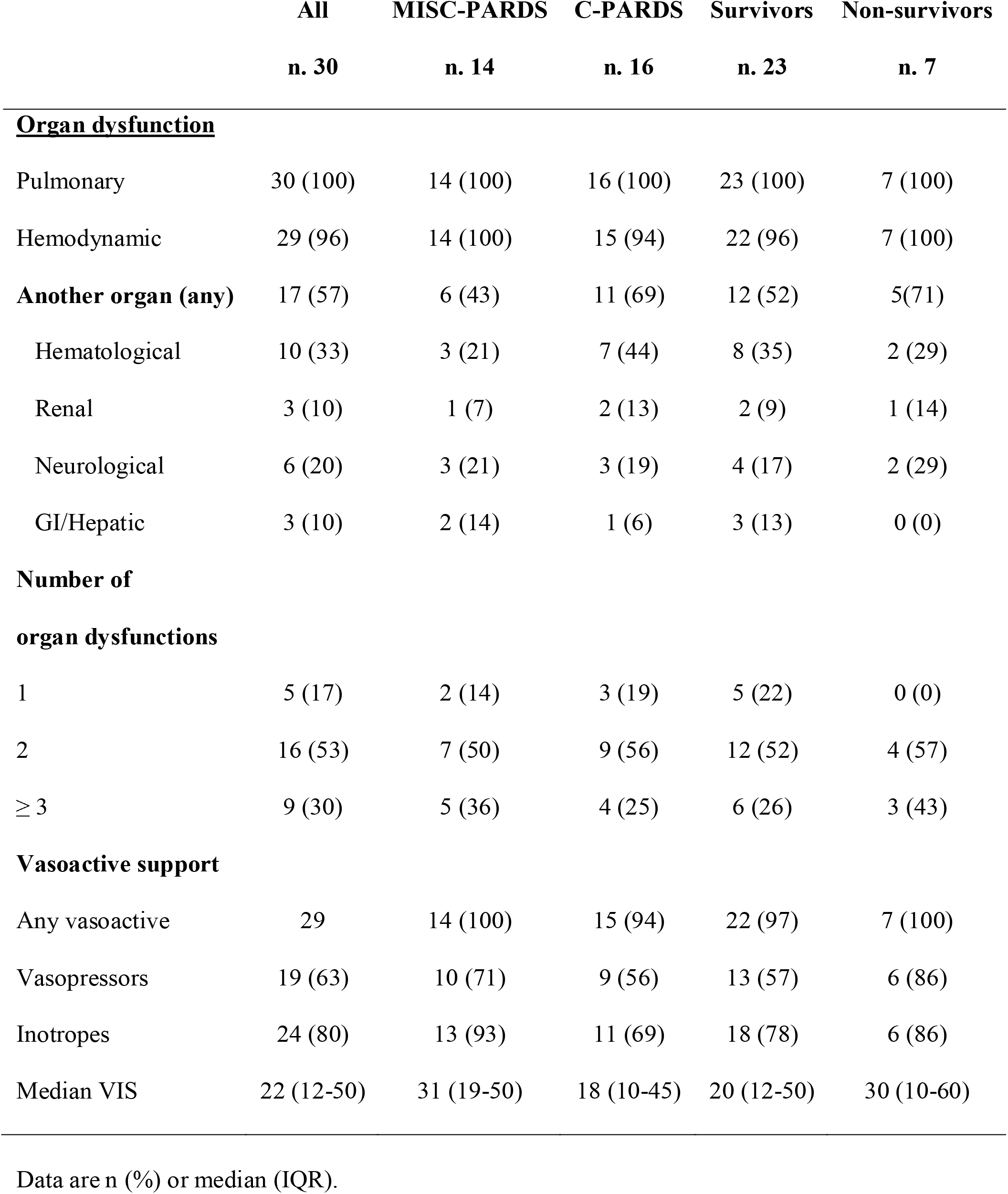
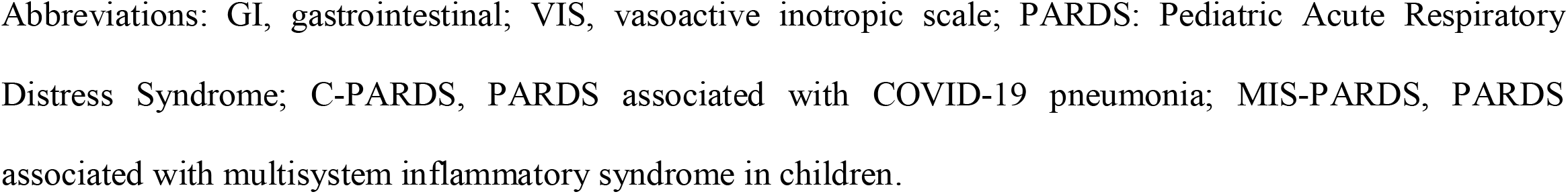
Organ failure and vasoactive support in pediatric respiratory distress syndrome associated with COVID-19 according to the clinical phenotypes and outcomes.

|  | All<br>n. 30 | MISC-PARDS<br>n. 14 | C-PARDS<br>n. 16 | Survivors<br>n. 23 | Non-survivors<br>n. 7 |
| --- | --- | --- | --- | --- | --- |
| **<u>Organ dysfunction</u>** |  |  |  |  |  |
| Pulmonary | 30 (100) | 14 (100) | 16 (100) | 23 (100) | 7 (100) |
| Hemodynamic | 29 (96) | 14 (100) | 15 (94) | 22 (96) | 7 (100) |
| **Another organ (any)** | 17 (57) | 6 (43) | 11 (69) | 12 (52) | 5(71) |
| Hematological | 10 (33) | 3 (21) | 7 (44) | 8 (35) | 2 (29) |
| Renal | 3 (10) | 1 (7) | 2 (13) | 2 (9) | 1 (14) |
| Neurological | 6 (20) | 3 (21) | 3 (19) | 4 (17) | 2 (29) |
| GI/Hepatic | 3 (10) | 2 (14) | 1 (6) | 3 (13) | 0 (0) |
| **Number of**<br>**organ dysfunctions** |  |  |  |  |  |
| 1 | 5 (17) | 2 (14) | 3 (19) | 5 (22) | 0 (0) |
| 2 | 16 (53) | 7 (50) | 9 (56) | 12 (52) | 4 (57) |
| ≥ 3 | 9 (30) | 5 (36) | 4 (25) | 6 (26) | 3 (43) |
| **Vasoactive support** |  |  |  |  |  |
| Any vasoactive | 29 | 14 (100) | 15 (94) | 22 (97) | 7 (100) |
| Vasopressors | 19 (63) | 10 (71) | 9 (56) | 13 (57) | 6 (86) |
| Inotropes | 24 (80) | 13 (93) | 11 (69) | 18 (78) | 6 (86) |
| Median VIS | 22 (12-50) | 31 (19-50) | 18 (10-45) | 20 (12-50) | 30 (10-60) |

Data are n (%) or median (IQR).
Abbreviations: GI, gastrointestinal; VIS, vasoactive inotropic scale; PARDS: Pediatric Acute Respiratory Distress Syndrome; C-PARDS, PARDS associated with COVID-19 pneumonia; MIS-PARDS, PARDS associated with multisystem inflammatory syndrome in children.

Fourteen patients (47%) were classified in MIS-PARDS phenotype and 16 (53%) children in C-PARDS phenotype. MIS-PARDS had more frequent abdominal pain and rash, while the C-PARDS group had more respiratory symptoms, like cough and lower oxygen saturation (Table 1). All patients with MIS-PARDS phenotype had a negative RT-PCR, and it was positive in 62.5% of the C-PARDS subgroup (p < 0.001). On the contrary, IgM(-)/IgG(+) profile was observed in 64.3% of MIS-PARDS and only in 16.7% of C-PARDS (p=0.02). (Table 1)

MIS-PARDS patients had significantly more cardiac dysfunction than the C-PARDS group (71 vs. 25%, p=0.03), and no other differences were found in organ failures and vasoactive support (Table 2). Mild left ventricular dysfunction was observed in 42% of MIS-PARDS and 6% of C-PARDS (supplementary table 2), but no severe dysfunction was found. C-PARDS group had more profound hypoxemia, higher Pplat and DP, and lower C_RS_, all p< 0.05, while MIS-PARDS had a higher PIP-Pplat gradient (p=0.04) (Table 3 and Figure 2). There was a trend toward higher mortality in the C-PARDS group compared to MIS-PARDS (38 vs. 7%, p=0.09). MV free days were significantly lower in C-PARDS than MIS-PARDS (p=0.02), without differences in other outcomes.

**Table 3.** Gas exchange, mechanical ventilation settings, and lung mechanics in pediatric respiratory distress syndrome associated with COVID-19 according to the clinical phenotypes and outcome.

|  | All<br>n. 30 | MISC-PARDS<br>n. 14 | C-PARDS<br>n. 16 | Survivors<br>n. 23 | Non-survivors<br>n. 7 |
| --- | --- | --- | --- | --- | --- |
| **<u>Oxygenation</u>** |  |  |  |  |  |
| FiO<sub>2</sub> | 60 (45-100) | 53 (40-60) | 60 (48-100) | 55 (45-60) | 100 (45-100) |
| PaO<sub>2</sub>, mmHg | 80 (60-100) | 92 (70-129) | 77 (55-83) | 85 (60-100) | 77 (56-80) |
| PaO<sub>2</sub>/FIO<sub>2</sub> | 130 (85-228) | 163 (129-252)* | 96 (74-150) | 130 (98-230) | 80 (70-171) |
| PaO<sub>2</sub>/FIO<sub>2</sub> < 100 | 12 (40) | 6 (38)* | 12 (86) | 15 (65) | 3 (43) |
| P(A-a) O<sub>2</sub>, mmHg | 246 (165-399) | 235 (141-305) | 336 (175-564) | 240 (141-330) | 547 (182-580) |
| Oxygenation index | 9.7 (6.1-19.9) | 7.5 (4.2-13.6)* | 11.9 (7.8-23.0) | 8.4 (4.3-15) | 22.5 (8.6-24) |
| PARDS severity |  |  |  |  |  |
| Mild (4 to <8) | 6 (20) | 4 (29) | 2 (13) | 6 (26) | 0 (0) |
| Moderate (8 to <16) | 15 (50) | 7 (50) | 8 (50) | 12 (52) | 3 (43) |
| Severe (≥ 16) | 9 (30) | 3 (21) | 6 (37) | 5 (22) | 4 (57) |
| **<u>Arterial Blood Gas</u>** |  |  |  |  |  |
| pH | 7.33 (7.19-7.39) | 7.30 (7.22-7.39) | 7.37 (7.2-7.44) | 7.35 (7.22-7.42) | 7.18 (6.9-7.39) |
| PaCO<sub>2</sub>, mmHg | 44 (33-51) | 38 (33-51) | 47 (34-56) | 38 (33-51) | 51 (45-59) |
| Bicarbonate, mmol/L | 21 (17-24) | 20 (17-23) | 22 (17-26) | 21 (18-24) | 17 (11-27) |
| Hemoglobin, g/dL | 10.3 (9.6-11.0) | 10.0 (10.0-11.0) | 10.6 (9.0-11.5) | 10.0 (9.6-12.0) | 10.6 (9.0-11.0) |
| Lactate, mmol/L | 1.6 (1.0-3.0) | 1.7 (1.2-3.4) | 1.4 (0.9-2.6) | 1.6 (0.9-3.0) | 2 (1.0-7.9) |
| **<u>Lung mechanics</u>** |  |  |  |  |  |
| PIP, cmH<sub>2</sub>O | 28 (24-32) | 27 (22-30) | 29 (25-33) | 25 (23-28)* | 33 (32-35) |
| Pplat | 20 (16-30) | 18 (15-20)* | 26 (19-30) | 18 (15-22)* | 30 (28-30) |
| PEEP, cmH<sub>2</sub>O | 7 (6-11) | 7 (5-10) | 8 (7-12) | 7 (6-11) | 10 (7-12) |
| Paw | 12.7 (11.0-16.4) | 11.5 (10.0-16.0) | 14.1 (11.6-17.5) | 12.2 (10.0-16.3) | 16 (12.1-18.3) |
| V<sub>T</sub>, ml/kg | 7 (6-8) | 7.0 (6.5-8.0) | 6.5 (6.0-7.5) | 7 (6-8) | 7 (6-9) |
| Prone Position | 9 (30) | 3 (21) | 6 (38) | 6 (26) | 3 (43) |
| Prone duration, hours | 48 (48-48) | 96 (48-96) | 48 (48-48) | 48 (48-48) | 48 (48-96) |
| Neuromuscular blocker | 13 (43) | 4 (29) | 9 (56) | 8 (35) | 5 (71) |

Data are n (%) or median (IQR); \*p<0.05 using Fisher's exact test or Mann-Whitney U test. Abbreviations: FiO<sub>2</sub>: Fraction of inspired oxygen; PaO<sub>2</sub>: Arterial oxygen partial pressure; PaO<sub>2</sub>/FIO<sub>2</sub>: PaO<sub>2</sub> to FiO<sub>2</sub> ratio; P(A-a) O<sub>2</sub>: Alveolar-arterial oxygen gradient; PEEP: Positive end-expiratory pressure; PaCO<sub>2</sub>: Partial pressure of carbon dioxide; PIP: Peak inspiratory pressure; Vt/Kg: Tidal volume per kilogram; Pplat: plateau pressure; Paw: mean airway pressure; C-ARDS, Coronavirus associated acute respiratory distress syndrome; PARDS: Pediatric Acute Respiratory Distress Syndrome; C-PARDS, PARDS associated with COVID-19 pneumonia; MIS-PARDS, PARDS associated with multisystem inflammatory syndrome in children

**Figure 2.**
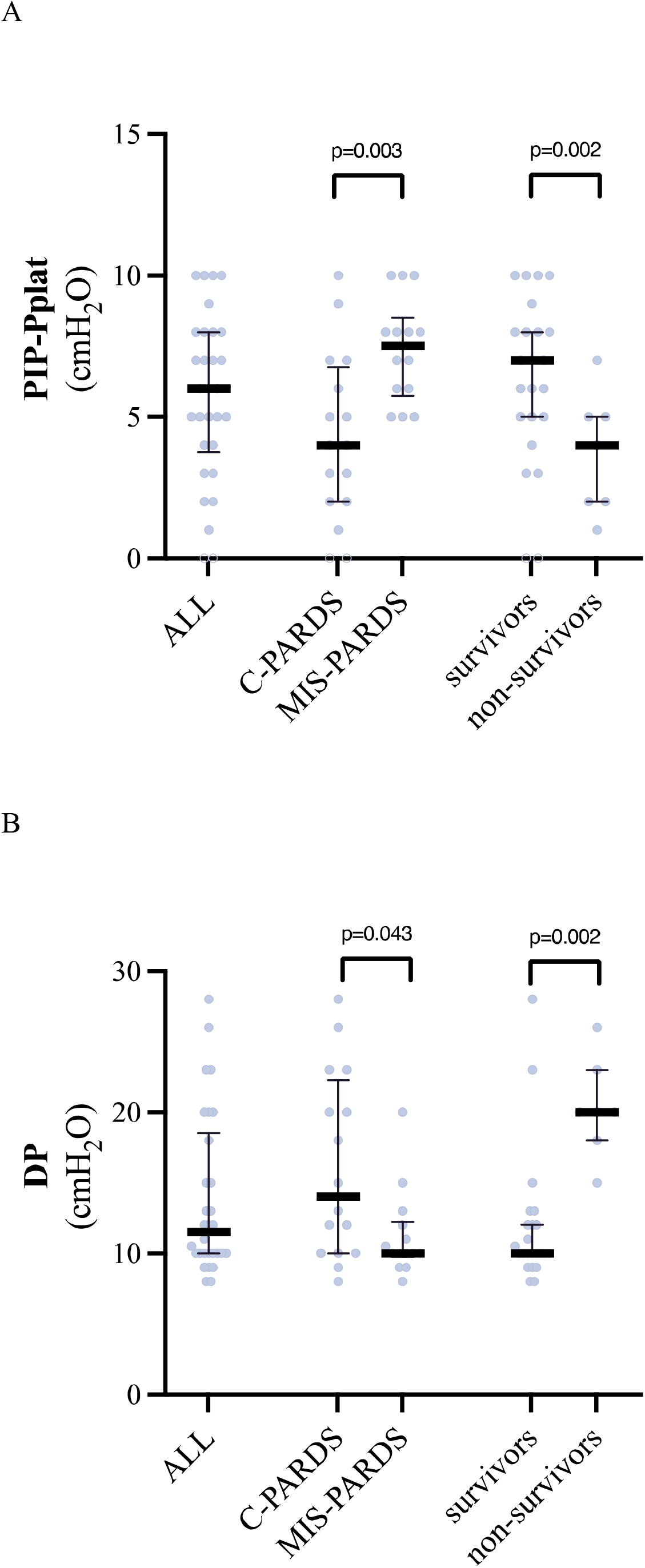
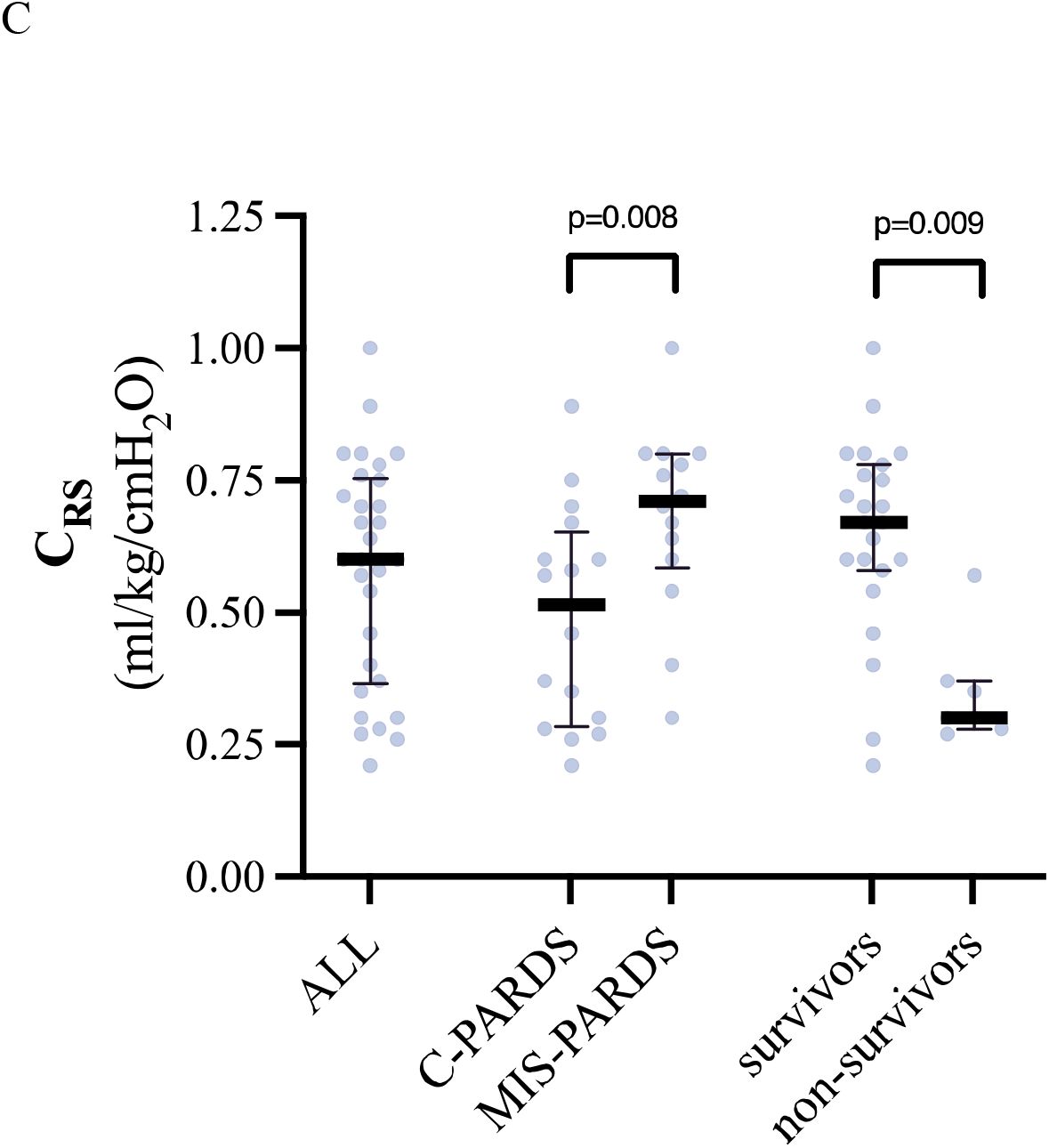
Boxplot graph of calculated parameters of lung mechanics of Children with PARDS associated with COVID-19, according to the clinical phenotypes and outcome. (A) The resistive component of work of breathing (Peak Inspiratory Pressure minus Plateau Pressure subtraction, [PIP-Pplat]); (B) the viscoelastic component of the work of breathing (Driving Pressure, DP); (C) compliance of the respiratory system (C_RS_). (PARDS: Pediatric Acute Respiratory Distress Syndrome; C-PARDS, PARDS associated with COVID-19 pneumonia; MIS-PARDS, PARDS associated with multisystem inflammatory syndrome in children)

When comparing survivors and non-survivors, there were no significant differences in demographics, organ dysfunctions, vasoactive support, and gas exchange. In addition, there were no significant differences in V_T_ and PEEP between both groups, but C_RS_ was significantly lower in non-survivors; thus, PIP, Pplat, and DP were higher (all p< 0.05).

ROC of DP and mortality showed an AUC of 0.91 (95% CI 0.81-1.00), and the best cut point was 15 cmH_2_O (100% sensitivity and 87% specificity). AUC for Elastance (1/CRS) was 0.89 (95% CI 0.77-1.00), with a best cut point of 2.7 (C_RS_ 0.37) (85.7% sensibility and 91.3% specificity). AUC for Pplat was 0.89 (95% CI 0.76-1.00), with a best-cut point of 28 (C_RS_ 0.37) (100% sensibility and specificity 87%). (Supplementary figure)

## DISCUSSION

The main result of this multicentric study of PARDS associated with pediatric critical COVID-19 was the description of two different phenotypes, C-PARDS and MIS-PARDS. C-PARDS is similar to other severe viral pneumonia, and we observed more frequent respiratory symptoms, hypoxia, and a positive COVID-19 rt-PCR in upper respiratory airways. On the other hand, MIS-PARDS had abdominal pain, mucocutaneous signs, and the characteristic MIS-C profile of IgG (+), IgM (-), and negative SARS-CoV-2 RT-PCR. In agreement with these observations, the lung mechanics parameters were different between subgroups: C-PARDS had higher Pplat and DP and lower C_RS_ and PIP-Pplat than MIS-PARDS. Regarding mortality, similar differences were observed between survivors and non-survivors. DP, Elastance (1/C_RS_), and Pplat were good discriminators for mortality, but DP had the best performance.

Remarkably, the C_RS_ of C-PARDS was close to the C_RS_ reported in other pediatric cohorts of viral PARDS^31^ and other restrictive lung diseases^38^. Regarding outcomes, mortality was not statistically different between C-PARDS and MIS-PARDS. VFD were significantly lower in this group, probably due to the primary pulmonary dysfunction observed in C-PARDS. Only one patient out of 14 died in the MIS-PARDS group, in accordance with the usual overall good prognosis of MIS-C.^11,12,15^

The mortality of our cohort may seem high, but when analyzing survival, many studies report between 10 and 70% of mortality in PARDS.^37^ Also, there are reports regarding differences in mortality between high and low-middle-income countries.^39,40^ It is very interesting that in the current study, previous known variables associated with mortality in PARDS and in pediatric critical COVID-19, like organ failures and hypoxemia severity, were not different between survivors and non-survivors.^37,38, 41-45^

A surprising result was the difference in the respiratory mechanics between the clinical phenotypes. The cohort fulfilled PARDS and critical COVID-19 criteria but with different characteristics. C-PARDS group’s features were like a classic moderate to severe primary ARDS. 46 On the contrary, the main characteristic of the MIS-PARDS group was an uncoupling between the respiratory mechanics and gas exchange. An attractive hypothesis is that the underlying pathophysiology leading to PARDS might be different in both phenotypes. In C-PARDS, airway epithelium and pneumocytes injury might be the initial alteration resulting in alveolar collapse with inflammatory exudate, neutrophils, and macrophages. On the other hand, in the context of systemic dysregulated inflammation, MIS-PARDS, the initial injury might depend on capillary endothelium, leading to interstitial edema and then flooding of alveolar space. It is important to recall severe left ventricular dysfunction was not observed in the cohort, but mild dysfunction was frequently reported, a common finding in critically ill children^26,47-49^. In addition, no signs of cardiogenic acute pulmonary edema were present.

A possible explanation for the trend toward higher mortality is the low C_RS_ of the C-PARDS group, resulting in higher DP, a previous parameter associated with mortality in ARDS patients. In an exploratory analysis, we found that DP was close to the ideal clinical discriminator for mortality. Interestingly, the best cut-off was 15 cmH_2_O. In 2 retrospective studies of children under MV due to acute hypoxemic respiratory failure, high DP (≥ 15 cmH_2_O) was associated with less VFD but not mortality. This threshold also was described in adults. In a meta-analysis that included nine prospective trials and more than 3,500 patients, Amato et al. showed that DP was the best variable correlated with survival, even in patients within the usual thresholds of a lung-protective MV strategy^50^. Even more, interventions that resulted in a decrease in DP were associated with a greater survival rate. Other authors have confirmed the association between DP and ARDS outcomes, and a threshold of 15 cmH_2_O has been incorporated into most lung-protective protocols.^51,52^

Our study has some limitations. We report respiratory mechanics in quasi-static conditions in VCV mode; thus, they cannot be extrapolated to other modes of MV with a deaccelerating flow, frequently used in pediatrics^44^. Presented data is the worst during the first 72h of admission, so time-dependent variables are not analyzed^43^. We did not investigate other parameters associated with PARDS severity or outcomes, i.e., dead space or mechanical power, because it was not part of our objective^53-55^. As in many multicenter studies, there might be differences in ventilatory strategy between sites, and comparisons were not possible given the heterogeneity of cases per center. The lack of consistency, especially PEEP titration, might influence some calculations of pulmonary mechanics. The small number of patients in each group might lead to type II statistical errors in some analyzed variables. There is also a risk of type I statistical errors, given the absence of statistical correction for multiple comparisons. Finally, the small number of cases probably influenced the lack of a statistical difference in mortality between C-PARDS and MIS-PARDS groups, although the difference was clinically relevant (38 vs. 7%). Nonetheless, we consider our results important to define high-risk groups of children with critical COVID-19 and as hypothesis-generating data for PARDS in the general PICU population.

## CONCLUSIONS

Our data show two different clinical phenotypes of pediatric critical COVID-19 PARDS with distinctive pulmonary mechanics features. C-PARDS group characteristics were like a classic moderate to severe primary ARDS. A decoupling between compliance and hypoxemia was more frequent in MIS-PARDS. Regarding outcomes, C-PARDS had less VFD and a trend toward higher mortality than MIS-PARDS. Data from the quasi-static calculations were associated with mortality; specifically, a DP ≥ 15 cmH2O was the best discriminator. Standardized pulmonary mechanics measurements in PARDS might reveal essential information to tailor the ventilatory strategy. Future studies may include these data for subgroup classifications and outcomes and ultimately improve the care of critically ill children.

## Data Availability

All data produced in this work are contained in the manuscript.

## Acknowledgments

None

## Authors’ contributions

Conceptualization and design: JDR, CMM, NAA. Data collection: JDR, YLD, PCV, CMR, MQC, MCA, GQF, MTP. Data curation: JDR, PVH, FD. Formal analysis and investigation: JDR, FD, PVH. Draft preparation: JDR, PVH, FD; Manuscript review and editing: JDR, PVH, FD, PC. JDR supervised the study at all stages. All authors read and approved the final version of the manuscript.

## Funding

No funding was involved in this research

## Availability of supporting data

The data supporting this study’s findings are available from the corresponding author, FD, upon reasonable request.

## Compliance with ethical standards

### Conflicts of interests

The authors declare no conflicts of interest.

## Ethical approval

Local IRB of each participating center approved the study (Approval letters 088-2021-CIEI-HNHU, 011-2021-CIEI-HEVES, 001-2021-COVID-HR, 42-IETSI-ESSALUD-2020).

## Informed consent

Informed consent was waived due to gathering anonymized data previously collected for administrative and benchmarking purposes.

## Supplementary files

**Table S1.**
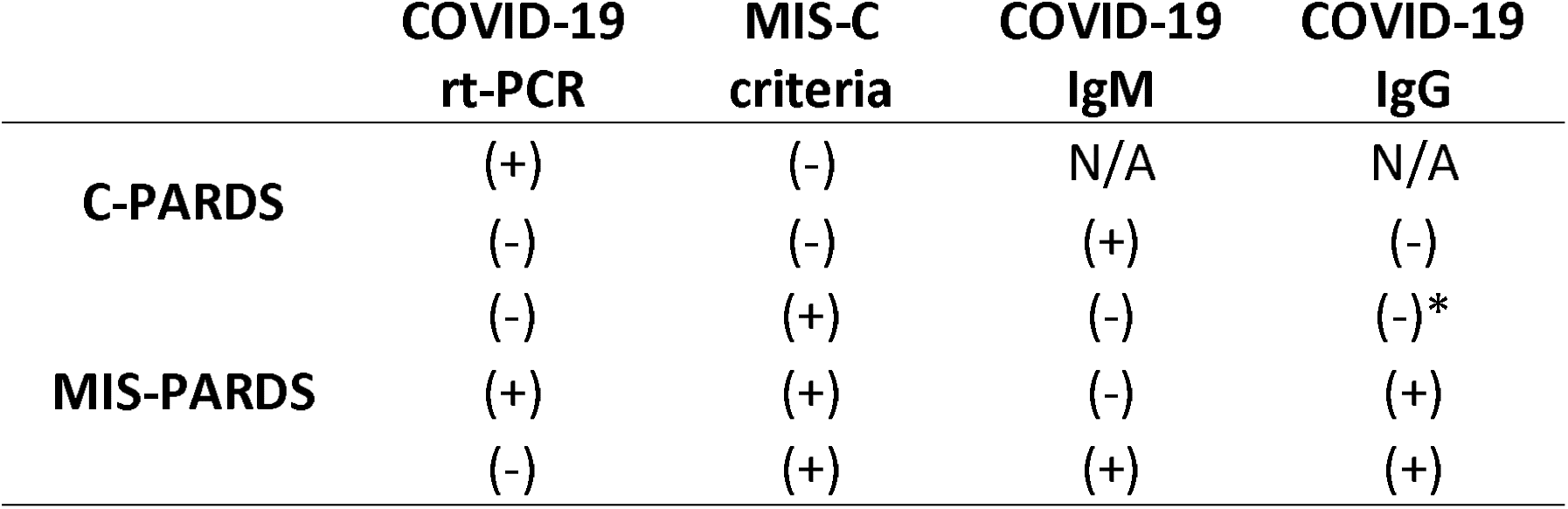
Case definitions for phenotypes according to diagnostics tests available. C-PARDS, pediatric acute respiratory syndrome associated to COVID-19 infection; MIS-PARDS, pediatric acute respiratory syndrome associated to multisystem inflammatory syndrome; rt-PCR, reverse transcription-polymerase chain reaction; MIS-C, multisystem inflammatory syndrome in children. * epidemiologic contact.

**Table S2.** Echocardiogram findings in C-PARDS and MIS-PARDS phenotypes. C-PARDS, pediatric acute respiratory syndrome associated to COVID-19 infection; MIS-PARDS, pediatric acute respiratory syndrome associated to multisystem inflammatory syndrome; LV, left ventricular.

|  | All<br>n = 30 | MIS-PARDS<br>n = 14 | C-PARDS<br>n = 16 |
| --- | --- | --- | --- |
| **Abnormal Coronaries** | 2 | 2 | 0 |
| **LV Dysfunction** | 4 | 4 | 1 |
| EF < 40% | 0 | 0 | 0 |
| **No Echocardiogram** | 10 | 1 | 9 |

**Figure S1.**
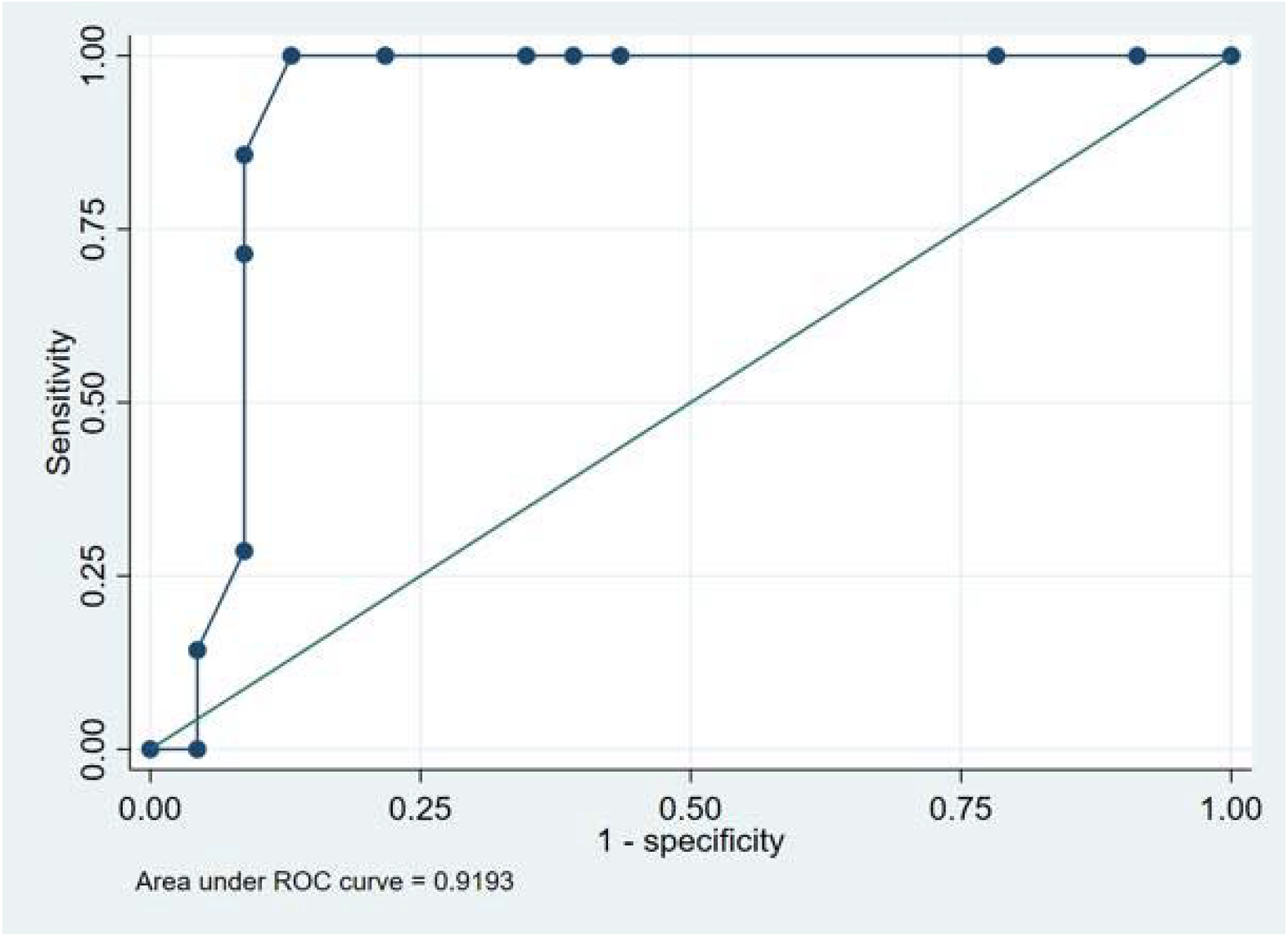
Receiver operator curve analysis for driving pressure and mortality. Area under the ROC curve was 0.91 (95% CI 0.81-1.00), and the best cut point was 15 cmH_2_O (100% sensitivity and 87% specificity).

## Notes

### Competing Interest Statement

The authors have declared no competing interest.

### Funding Statement

This study did not receive any funding

### Author Declarations

Approval letters 088-2021-CIEI-HNHU Hospital Nacional Hipolito Unanue, 011-2021-CIEI-HEVES Hospital de Emergencias de Villa El Salvador, 001-2021-COVID-HR Hospital Regional de Cusco, 42-IETSI-ESSALUD-2020 Hospital Edgardo Rebagliati Martins

